# Effect of Temporal Trend in Inflammatory and Cholesterol Risk on the Prognosis of Percutaneous Coronary Intervention-Treated Patients with Contemporary Statin Therapy

**DOI:** 10.1101/2023.10.17.23297180

**Authors:** Ang Gao, Zifeng Qiu, Yong Wang, Tingting Guo, Yanan Gao, Qianhong Lu, Zhiqiang Yang, Zhifan Li, Hong Qiu, Runlin Gao

**Author notes:** Corresponding author: Hong Qiu MD, PhD, Department of Cardiology, Cardio-Metabolic Medicine Center, Fuwai Hospital, National Center for Cardiovascular Diseases, Chinese Academy of Medical Sciences and Peking Union Medical College, Beijing, China. Ang Gao and Zifeng Qiu contributed equally to the article.

## Abstract

**Background:** Atherosclerotic cardiovascular disease patients still suffer from recurrent vascular events due to residual cholesterol and inflammatory risk. However, the relative importance of inflammation and cholesterol risk might have changed in percutaneous coronary intervention (PCI)-treated patients after accepting contemporary statin therapy. Hence, this study aims to evaluate the effect of temporal trend in inflammatory and cholesterol risk on the prognosis of that population.

**Methods:** PCI-treated patients at Fuwai Hospital between 1^st^ January 2016 and 31^st^ December 2017 with on-admission and follow-up high-sensitive C-reactive protein (hs-CRP) and low-density lipoprotein cholesterol (LDL-C) within 1 to 3 months were retrospectively enrolled. Participants were all taking contemporary statin treatment at discharge. Tertiles of on-admission and follow-up hs-CRP (a biomarker for inflammatory risk) and LDL-C (a biomarker for cholesterol risk) were assessed as determinants of one-year major adverse cardiovascular and cerebrovascular events (MACCEs). Multivariable Cox proportional hazard model was used to evaluate the prognostic value of on-admission, follow-up cholesterol and inflammatory risk. High inflammatory or cholesterol risk after accepting contemporary statins were expressed as residual cholesterol risk (RCR), residual inflammatory risk (RIR) and residual cholesterol and inflammatory risk (RCIR). Subgroup analysis of inflammatory and cholesterol risk on admission was conducted based on the glycometabolic status, index presentation and guideline-recommended statin therapy (GRST) at discharge.

**Results:** After one-year of follow-up, 187 MACCEs occurred in 2373 participants. Among the on-admission and follow-up hsCRP and LDL-C tetiles, only the follow-up LDL-C tertile failed to predict the occurrence of MACCEs [T3 versus T1, adjusted hazard ratio (HR) 0.89, 95% confidence interval (CI) 0.61-1.29, *P*=0.544]. After adjusting for various confounding factors, on-admission high cholesterol and inflammatory risk was significantly associated with the incidence of MACCEs (HR 2.45 95%CI 2.45 1.42-4.21, *P*<0.001). RIR can be a major determinant of MACCEs (adjusted HR 4.43, 95% CI 2.82-6.98, *P*<0.001). Subgroup analysis showed the potential predictive role of on-admission high inflammatory risk only for MACCEs in those with diabetes mellitus (HR 2.35, 95% CI 1.01-5.43) and accepting underpowered statins at discharge (HR 2.16, 95% CI 1.05-4.41).

**Conclusion:** We observed a combined effect of on-admission high cholesterol and inflammatory risk that could predict the risk of MACCEs. Inflammatory risk assessed by hs-CRP was a stronger predictor for MACCEs than cholesterol risk assessed by LDL-C in PCI-treated patients after taking contemporary statin therapy. Additionally, on-admission high inflammatory risk only could independently predict cardiovascular outcomes in PCI-treated patients with diabetes mellitus and accepting underpowered statin therapy.

## Introduction

Owing to the aging population and the increasing prevalence of cardiometabolic risk factors, cardiovascular deaths have become the leading cause of mortality in China[1]. Statins has been recognized as the cornerstone of cardiovascular secondary prevention due to their effectiveness in lowering the rates of recurrent myocardial infarction, stroke and cardiovascular deaths, which has been substantiated in previous randomized controlled trials[2]. However, statin-treated patients, especially those with advanced atherosclerotic cardiovascular disease (ASCVD), still suffer from a relatively high incidence of MACCEs even after an early revascularization strategy, an issue commonly ascribed to the problem of ‘residual risk’[3]. RCR, defined as an unachieved LDL-C goal in clinical practice despite contemporary statin treatment, undoubtedly represents a residual risk factor and provides evidence for large-scale clinical trials targeting more aggressive lipid-lowering therapies. In the IMPROVE-IT trial, Ezetimibe, a non-statin drug that selectively inhibits intestinal absorption of cholesterol, which could effectively reduce LDL-C concentrations by 25% on the basis of statins, demonstrated a 2% absolute risk reduction of the clinical endpoints among myocardial infarction patients [4, 5]. Proprotein convertase subtilisin/kexin type 9 (PCSK9) inhibitor, known for significantly reduction of LDL-C by 55-75%, showed a 15% risk reduction in the primary endpoint in the FOURIER and ODYSSEY OUTCOMES trials[6, 7]. Another major determinant of residual risk is inflammation, which contributed with similar magnitude as lipids to the atherosclerotic process and served as a better predictor of cardiovascular outcomes than LDL-C under current statin treatment[8]. Residual inflammatory risk, defined as unachieved hs-CRP goal (≥2 mg/L) despite LDL-C<1.8 mmol/L under optimal treatment. Persistently high inflammatory risk exists across Western and East Asian populations after percutaneous coronary intervention (PCI) treatment and could predict the incidence of MACCEs even in individuals with achieved baseline LDL-C levels[9–11]. Recent RCTs targeting anti-inflammatory agents also produced positive results. Colchicine, as a generic anti-inflammatory agent, could effectively lower recurrent cardiovascular event risk in those with established ASCVD[12, 13]. Recently, a study focusing on real-world population found that patients with cardiovascular diseases and high residual cholesterol and inflammatory risk had an increased risk of all-cause mortality[14]. However, there’s lack of measurements of hs-CRP and LDL-C level during follow-up and the relative importance of inflammatory risk and cholesterol risk after taking statin treatment remained elusive. Besides, the inflammatory level, especially in unstable patients, is easily affected by the index presentation of PCI, and the cholesterol burden could be alleviated after current pharmacological strategies. Serial measurements of inflammatory and cholesterol levels could better explain the inconsistencies surrounding hs-CRP and LDL-C in previous clinical studies. There is lack of studies concentrating on serial measurements of hs-CRP and LDL-C to understand the dynamic changes in RCR and RIR in PCI-treated individuals receiving contemporary statin treatment. Hence, the aim of this study was to evaluate the effect of the dynamic change of the inflammatory and cholesterol risk on the prognosis of PCI-treated patients with contemporary statin therapy.

## Methods

### Study design and population

A total of 2373 consecutive PCI-treated patients at Fuwai Hospital between 1^st^ January 2016 and 31^st^ December 2017 with serial measurements of hs-CRP and LDL-C (on admission and within 1 to 3 months after PCI treatment) were recruited into this single-center, retrospective observational study. This study was in accordance with the principles of the Declaration of Helsinki, and all patients signed informed consent before discharge. The detailed enrolment procedure is depicted in Fig 1. The exclusion criteria included the following: 1) failure to complete the 12-month follow-up; 2) major adverse clinical events within 3 months after the PCI procedure; 3) infectious diseases, malignant tumors, immune system disorders or suspected familial hypercholesterolemia (LDL-C≥4.7 mmol/L); and 4) inability to accept statin therapy for various causes at discharge. Hs-CRP and LDL-C were measured at least twice (on admission and 1 to 3-month follow-up); if one or more hs-CRP and LDL-C values were measured during the 1 to 3-month follow-up, the highest value was incorporated into the analysis. Participants were stratified into 4 groups according to on-admission hs-CRP (a biomarker for inflammatory risk) and LDL-C (a biomarker for cholesterol risk) levels: 1) participants with no cholesterol and inflammatory risk (hs-CRP< 2 mg/L and LDL-C<1.8, the reference group); 2) participants with high cholesterol risk only (hs-CRP<2 mg/L and LDL-C≥1.8 mmol/L); 3) participants with high inflammatory risk only (hs-CRP≥2 mg/L and LDL-C<1.8 mmol/L); and 4) participants with high cholesterol and inflammatory risk (hs-CRP≥2mg/L and LDL-C≥1.8mmol/L). Participants with follow-up cholesterol and inflammatory risk after accepting contemporary statins were further expressed as participants with no residual risk, participants with RCR, participants with RIR and participants with RCIR.

**Fig 1.**
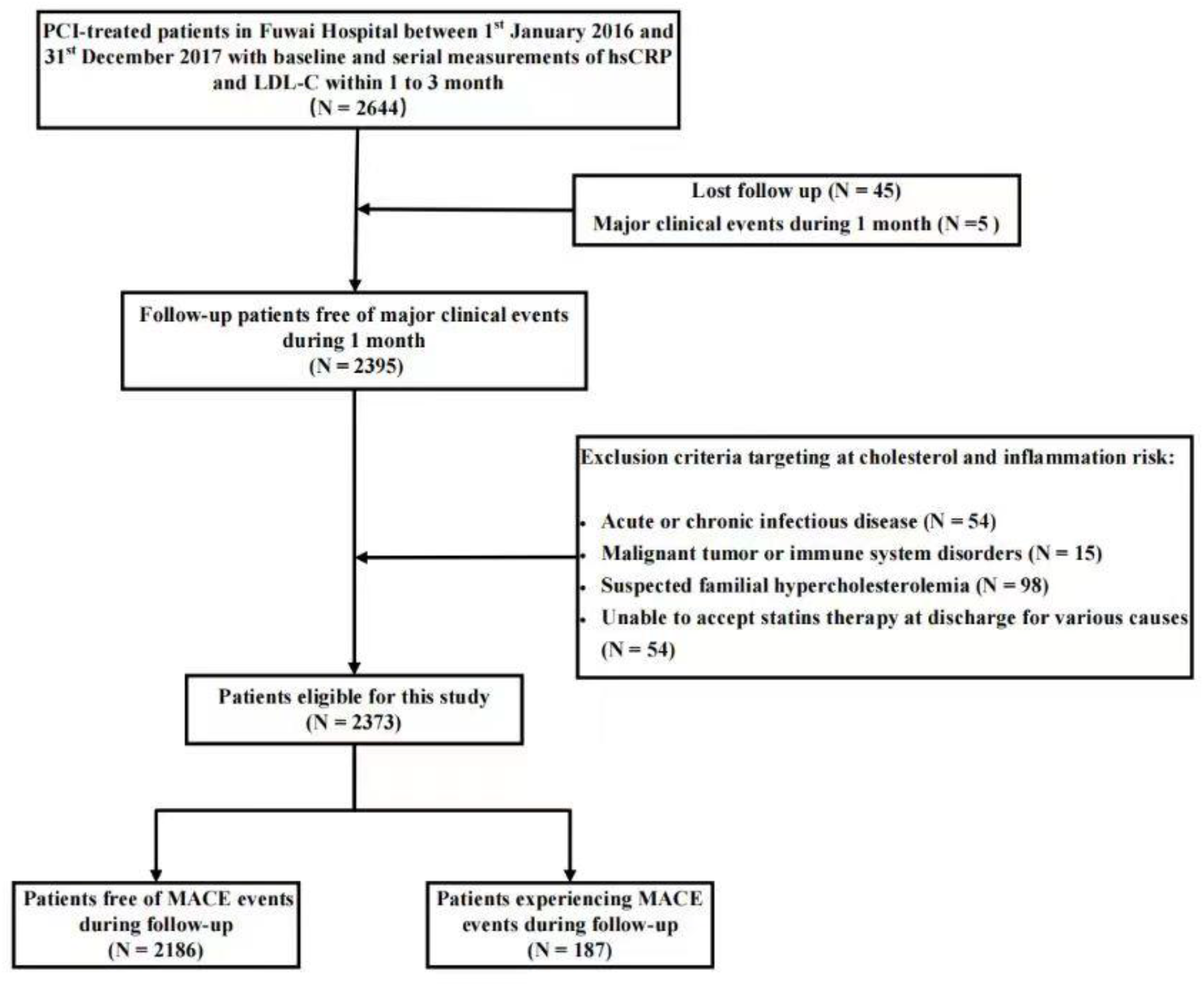
Flow diagram of patient selection

### Clinical outcomes

The primary endpoint of this study was the occurrence of MACCEs after PCI, defined as the composite of all-cause death, acute myocardial infarction, any unplanned coronary revascularization, rehospitalization for other cardiovascular causes, or cerebrovascular events comprising stroke or transient ischemic attack.

### Data collection and disease definition

Demographic data and medication at discharge were obtained through a review of medical records, which was approved by Fuwai Hospital. On-admission biochemical assessment was performed in whole blood samples drawn after arrival to the emergency or general ward. Blood samples were taken after overnight fasting if participants were not indicated for emergent coronary revascularization. At the 1 to 3-month visits, follow-up biochemical measurements were all calculated using blood samples taken from the vein after overnight fasting. Laboratory indicators, including hs-CRP and LDL-C, were all measured by standard techniques. All enrolled participants were followed up for 12 months or until the time of a major adverse clinical event. Follow-up was performed by telephone interviewers using standardized questionnaires at 6 and 12 months after the PCI treatment. The intensity of statins was defined according to ACC/AHA guideline definitions[15]. High-intensity statin in current study included: Atorvastatin 40 mg, Rosuvastatin 20 mg; moderate intensity statin included: Atorvastatin 10-20 mg, Rosuvastatin 5-10 mg, Pravastatin 40 mg, Simvastatin 20 mg, Fluvastatin 80 mg or Pitavastatin 4 mg; low intensity statin included: Pravastatin 20 mg, Fluvastatin 40 mg or Pitavastatin 1-2 mg. Guideline recommended statin treatment (GRST) was defined as the prescription of high-intensity statins in patients ≤75 years or a moderate-intensity statin or high-intensity statin in patients >75 years. The diagnosis of diabetes mellitus was based on the previous diagnosis and treatment with glucose-lowering medication or recommendations from the American Diabetes Association[16]. Hypertension was defined by the recommendations from the European Society of Hypertension, an office systolic blood pressure value≥140 mmHg or a diastolic blood pressure value≥90 mmHg or the use of antihypertensive drugs in the past 2 weeks[17]. Dyslipidemia was characterized by an increased total cholesterol, LDL-C or triglyceride level or a decreased high-density lipoprotein cholesterol level according to the third report of the National Cholesterol Education Program[18]. Acute myocardial infarction was defined as increased cardiac troponin values with ischemic symptoms or ischemic changes on electrocardiogram or imaging evidence of recent loss of viable myocardium or new regional wall motion abnormalities that were not related to the procedure[19]. The SYNTAX scores were calculated according to the preprocedural angiograms using the online calculation tool. Only coronary lesion≥50% stenosis in vessel ≥1.5mm was regarded as a positive lesion and was included into the calculation tool. The residual SYNTAX SCORE was calculated based on the remaining untreated obstructive coronary diseases after PCI treatment. If patients accepted staged PCI procedures, then the residual SYNTAX score after the last revascularization would be incorporated into the analysis.

### Statistical analysis

To better understand the characteristics of PCI-treated patients with different cholesterol and inflammatory burdens, participants were categorized into four groups according to on-admission and follow-up cholesterol and inflammatory risks. Descriptive variables are expressed as the mean ±standard deviation or median with interquartile range. Categorical variables are presented as frequencies and percentages, and the differences between no residual risk, RIR, RCR and RCIR group were determined by the one-way analysis of variance or the Kruskal-Wallis H test for normally or nonnormally distributed variables. Differences in categorical variables between the groups were compared by the Fisher’s exact or χ^2^ test for categorical variables. The results were shown in Table 1. The differences between no risk, inflammatory risk only, cholesterol risk only and high cholesterol and inflammatory risk groups were shown in Additional file 1: Table S1. Differences between the MACCE group and the survival group were compared by Student’s t test or the Mann-Whiteney U test for normally or nonnormally distributed continuous variates. The results were shown in Additional file 2: Table S2. Cumulative event rates were compared using the log-rank test, and the Kaplan-Meier method was used to depict the time-to-event curves. Univariable Cox proportional hazard analysis was performed to preliminarily explore the HRs for MACCEs among inflammatory and cholesterol risk groups. The association between cholesterol risk and inflammatory risk and MACCEs was finally determined using a multivariable Cox regression model after adjusting for the following confounders derived from univariable Cox proportional hazard model: hypertension, diabetes mellitus, dyslipidemia, body mass index (BMI), previous PCI, coronary artery bypass grafting (CABG) and myocardial infarction, left ventricular ejection fraction (LVEF), antiplatelet therapy, angiotensin blockade, diuretic and spironolactone use, multivessel disease, target vessel length, SYNTAX score and residual SYNTAX score. A two-sided *P* value<0.05 was considered statistically significant. All statistical analyzes were conducted using SPSS 23.0 (IBM SPSS statistics).

**Table 1.**
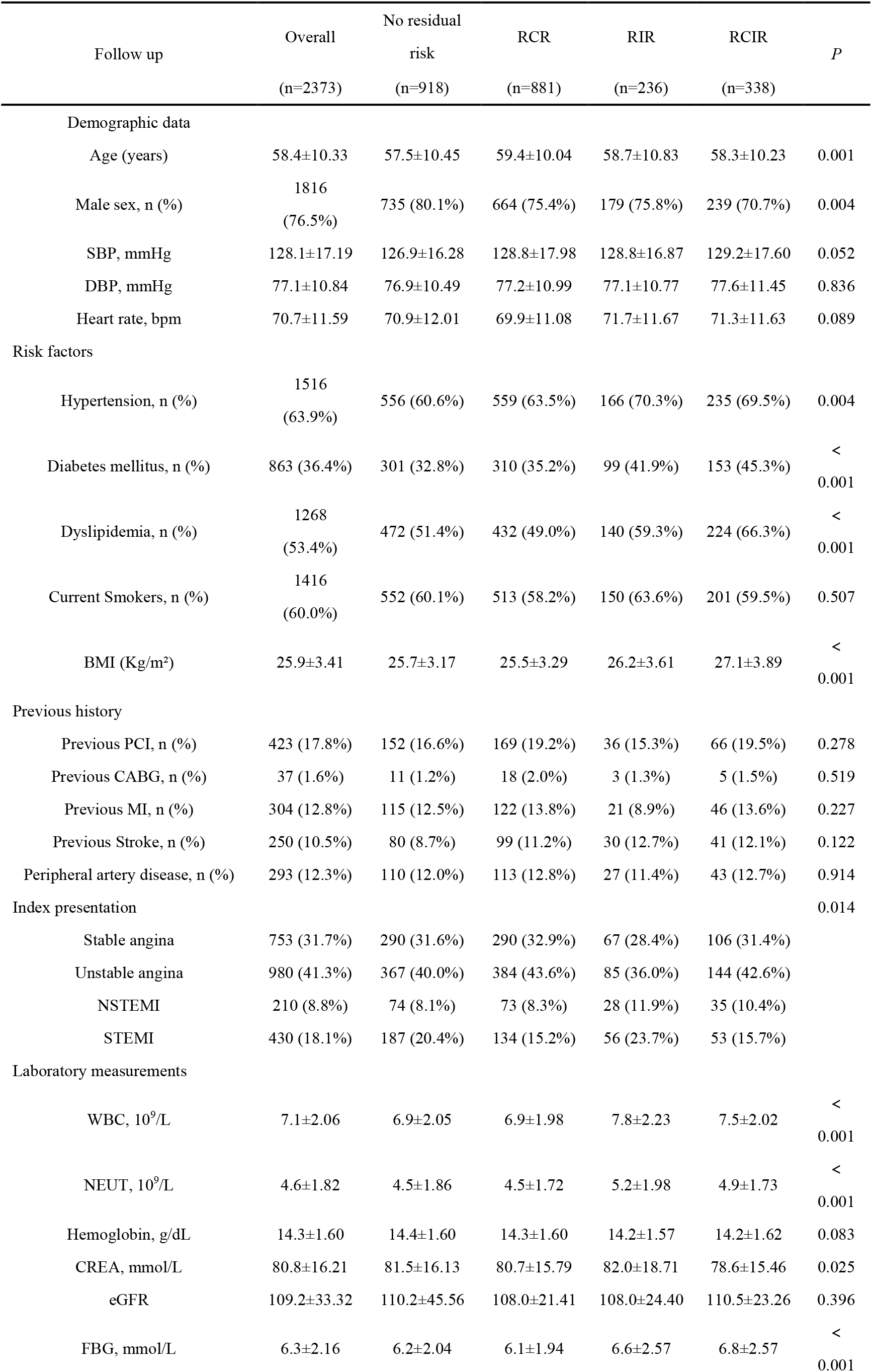
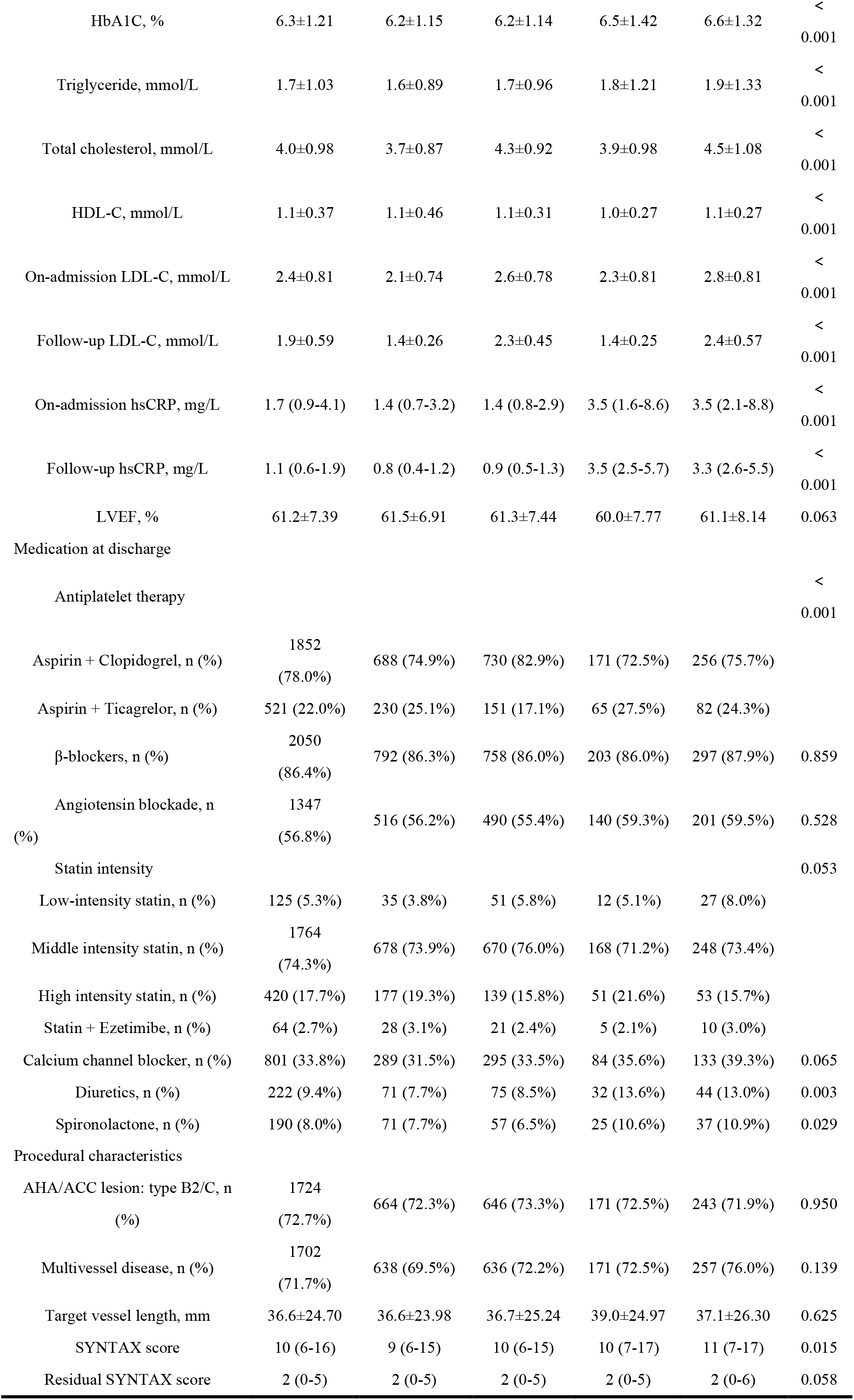
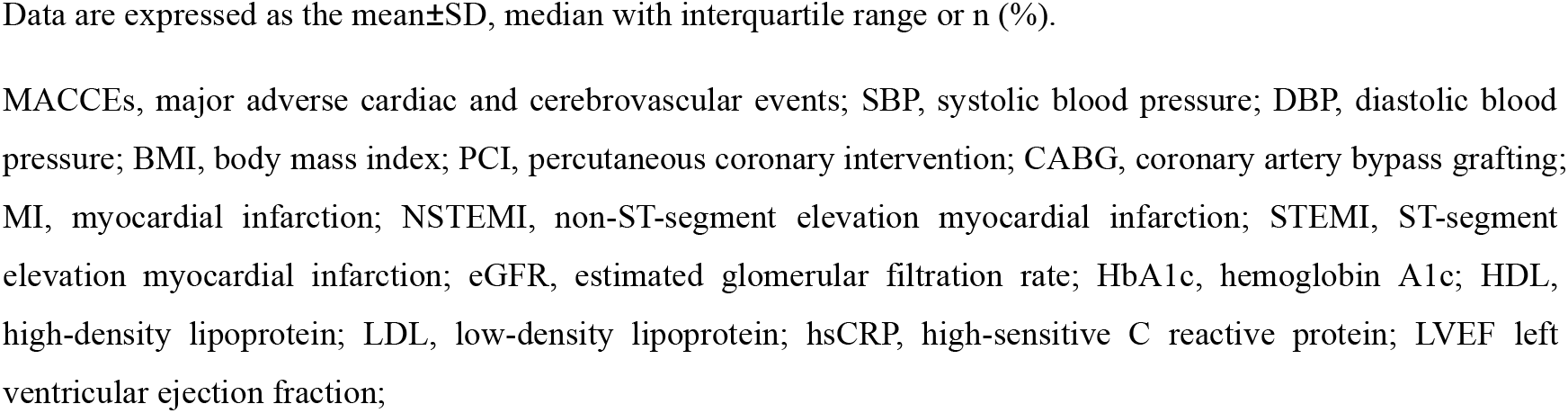
Baseline characteristics of PCI-treated patients stratified by follow-up inflammatory and cholesterol risk.

## Results

### Baseline characteristics

Fig 1 depicted the detailed enrollment procedure of the current study. Of the 2373 PCI-treated patients with serial measurements of hsCRP and LDL-C, 187 individuals experienced MACCEs during the12-month follow-up. The distribution of inflammatory and cholesterol risk on admission and follow-up were presented in Fig 2. Fig 2A and B showed that the share of patients with RCIR greatly decreased after contemporary statin treatment, decreasing from 36.6% to 14.2%, but the distribution of RCR and RIR remained almost unchanged after contemporary statin treatment. Fig 2C showed the share of patients from the perspective of persistent inflammatory risk. Almost one-fifth of patients (18.0%) were categorized into the persistently high inflammatory risk group (with on-admission and follow-up hsCRP≥2 mg/L) despite contemporary statin therapy. The burden of persistent cholesterol risk was shown in Fig 2D. A total of 45.2% of PCI-treated patients in the current study still experienced persistently high cholesterol risk (with on-admission and follow-up LDL-C≥1.8 mmol/L). The baseline characteristics of participants according to on-admission and follow-up cholesterol and inflammatory risk were shown in Additional file 1 Table: S1 and Table 1. The differences between participants with MACCEs versus those free of MACCEs during follow-up are presented in Additional file 2 Table S2. Of the 2373 enrolled participants, 1618 (81.2%) patients presented with acute coronary syndrome on admission. No differences were found in age and sex composition between the MACCE and survival groups. Participants experiencing MACCEs during follow-up had higher BMI, more rates of cardiovascular risk factors, such as diabetes mellitus, dyslipidemia, and hypertension, and were more likely to accept advanced antiplatelet strategies (aspirin + ticagrelor) and therapies targeting heart failure, such as angiotensin blockade, spironolactone and diuretics, and had a lower LVEF. The proportions of previous myocardial infarction, PCI and CABG were significantly higher in those with MACCEs. As with anatomical differences between the two groups, the MACCE group had higher SYNTAX and residual SYNTAX scores, longer target vessel lengths and higher rates of multivessel diseases.

**Fig 2.**
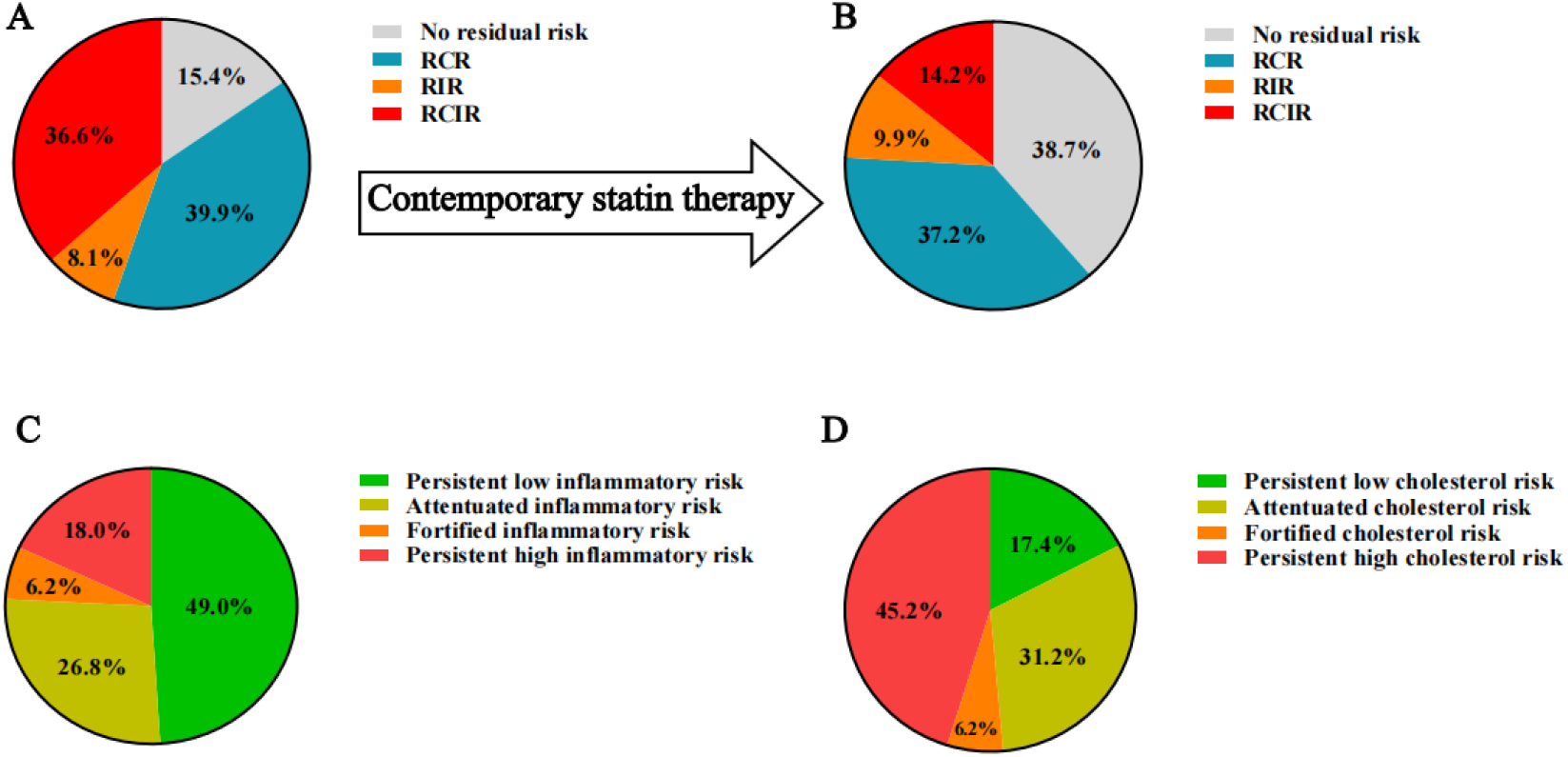
The distribution of PCI-treated patients regarding residual inflammatory and cholesterol risk burden (as assessed by high-sensitive CRP and LDL-C) on admission (A) and after contemporary statin therapy (B). The share of constitute inflammatory risk among PCI-treated patients stratified by high-sensitive CRP≥2mg/L criteria (C). The share of constitute cholesterol risk among PCI-treated patients stratified by LDL-C≥1.8mmol/L criteria (D). PCI, percutaneous coronary intervention; CRP, C reactive protein; LDL-C, low density lipoprotein cholesterol; RCR, residual cholesterol risk; RIR residual inflammatory risk; RCIR residual cholesterol and inflammatory risk

### Clinical outcomes according to on-admission and follow-up inflammatory and cholesterol levels

To preliminarily explore the predictive value of hs-CRP and LDL-C for one-year MACCEs, participants were first divided into 3 tertiles according to on-admission and follow-up hs-CRP and LDL-C levels. During the follow-up period, Kaplan-Meier curves showed differences in the risk of MACCEs between tertiles of on-admission and follow-up hs-CRP (all *P*<0.001) and on-admission LDL-C (*P*=0.018), except for the tertile of follow-up LDL-C (*P*=0.530) (Fig. 3 and Table 2). To further understand the impact of temporal trend in residual inflammatory and cholesterol risk on MACCEs, patients were stratified into on-admission and follow-up high cholesterol and inflammatory groups. Compared with the no cholesterol and inflammatory risk group as a reference, the HRs (95% CIs) of on-admission high cholesterol risk only, high inflammatory risk only and high cholesterol and inflammatory risk for MACCEs were 1.21 (0.68-2.15), 1.97 (0.99-3.95), and 2.45 (1.42-4.21) after adjusting for confounding factors. A significant association was found only between on-admission high cholesterol and inflammatory risk and MACCEs, as shown in Fig 4A and B and Table 3. After contemporary statin therapy, follow-up RIR and RCIR were both significantly associated with the risk of MACCEs, with HRs (95% CI) of 4.43 (2.82-6.98) and 4.23 (2.79-6.41), respectively (Fig 4C and D and Table 3). Fig 5 showed the predictive value of on-admission high cholesterol risk only, high inflammatory risk only and high cholesterol and cholesterol risk for the risk of MACCEs in different subgroups when stratified by glycometabolic status, index presentation and prescription of GRST at discharge. The predictive value of on-admission high cholesterol and inflammatory risk remained persistent across all subgroups. An association was found between high inflammatory risk only and MACCEs in the diabetes mellitus and underpowered statin groups, indicating the additional prognostic value of high inflammatory risk for MACCEs in this population.

**Fig 3.**
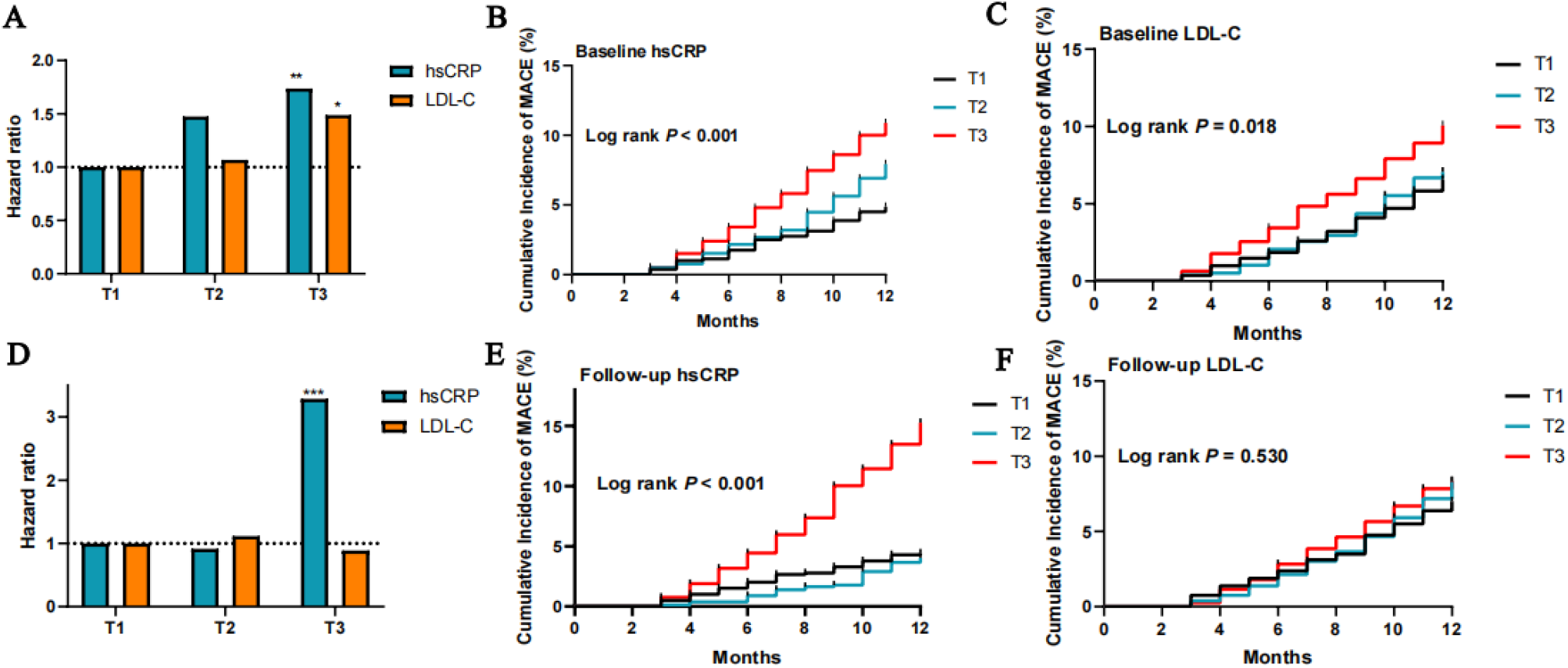
The hazard ratios for one-year MACCEs according to the tertile of baseline and follow-up high-sensitive CRP and LDL-C level (Fig 3A and 3D). Kaplan-Meier curves for one-year MACCEs based on the tertile of baseline and follow-up high-sensitive CRP (Fig.3B and 3C) and LDL-cholesterol (Fig.3E and 3F). MACCEs Major adverse cardiovascular and cerebrovascular events; CRP C-reactive protein; LDL-C Low density lipoprotein cholesterol; *P<0.05, **P<0.01, ***P<0.001

**Fig 4.**
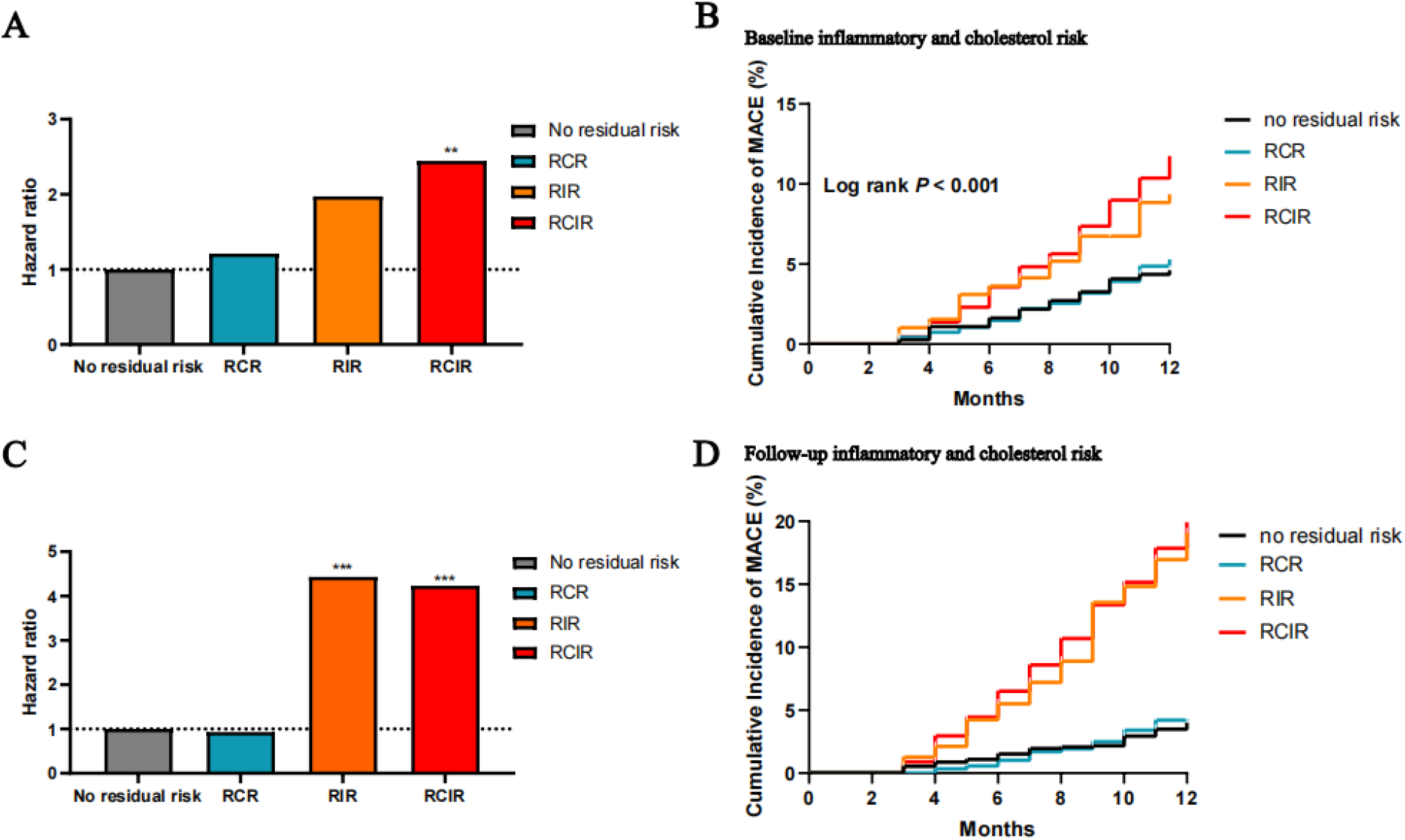
The hazard ratios for one-year MACCEs according to baseline and follow-up residual inflammatory and cholesterol risk (Fig 4A and 4C). Kaplan-Meier curves for one-year MACCEs based on baseline and follow-up residual inflammatory and cholesterol risk (Fig 4B and 4D). MACCEs, major adverse cardiac and cerebrovascular events; RCR residual cholesterol risk; RIR residual inflammatory risk; RCIR residual cholesterol and inflammatory risk; *P<0.05, **P<0.01, ***P<0.001

**Fig. 5.**
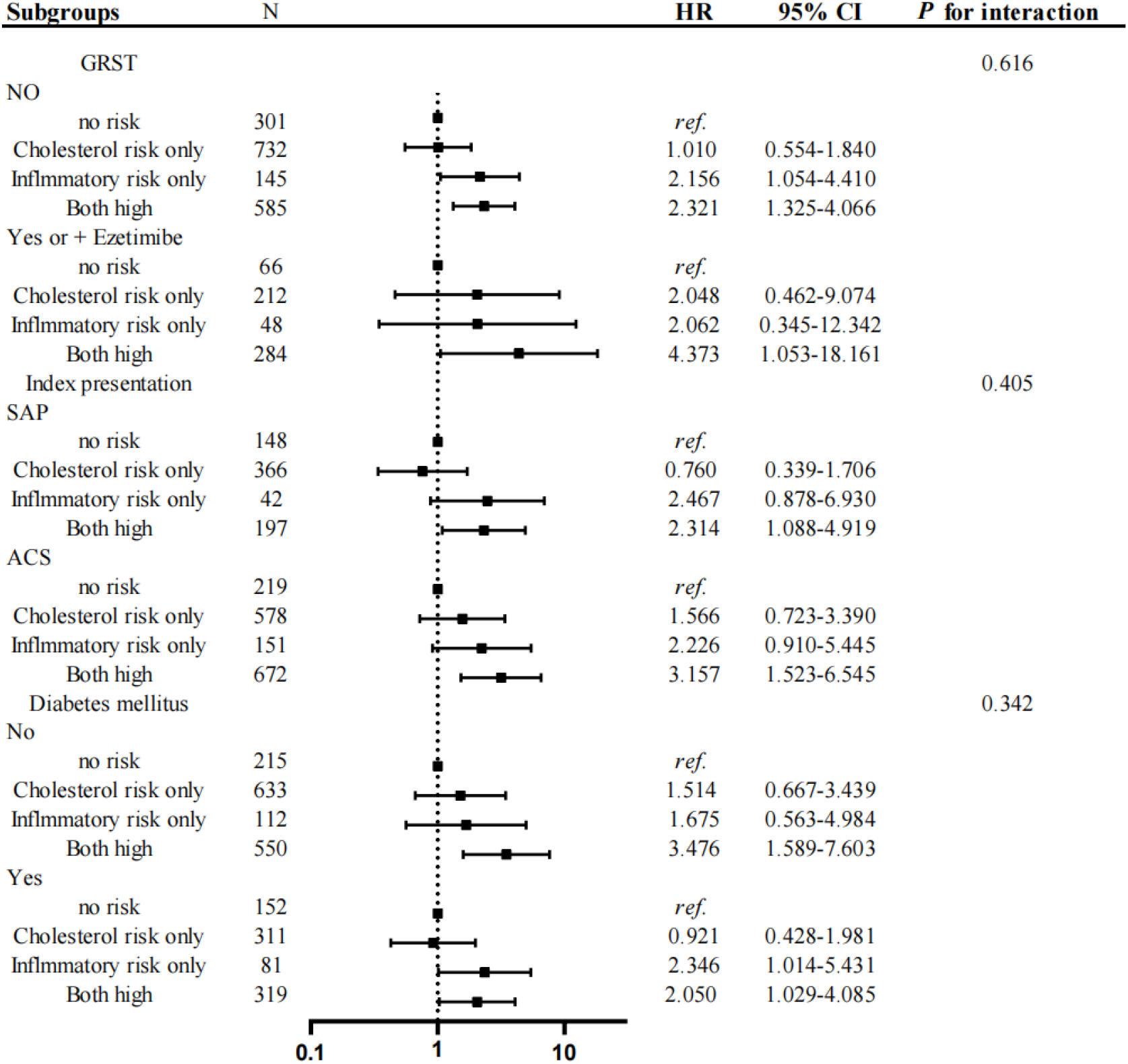
Comparative hazard ratios of RCR, RIR and RCIR for MACCEs across subgroups. MACCEs, major adverse cardiovascular and cerebrovascular events; GRST, guideline-recommended statin therapy; SAP, stable angina pectoris; ACS, acute coronary syndrome;

**Table 2.** The predictive value for one-year MACCEs according to the tertile of baseline and follow-up hsCRP and LDL-C levels.

| Baseline hsCRP | Events<br>n/N | Unadjusted Model |  |  | Adjusted Model |  |  |
| --- | --- | --- | --- | --- | --- | --- | --- |
|  |  | HR | 95% CI | P Value | HR | 95% CI | P Value |
| T1 (T1≤1.12) | 39/800 | 1 | Ref. |  | 1 | Ref. |  |
| T2 (1.13≤ T2≤2.97) | 62/782 | 1.642 | 1.100-2.451 | 0.015 | 1.475 | 0.971-2.242 | 0.068 |
| T3 (T3 > 2.97) | 86/791 | 2.299 | 1.575-3.357 | <0.001 | 1.736 | 1.147-2.629 | 0.009 |
| Baseline LDL-C |  |  |  |  |  |  |  |
| T1 (T1≤2.00) | 53/810 | 1 | Ref. |  | 1 | Ref. |  |
| T2 (2.01≤ T2≤2.70) | 55/779 | 1.078 | 0.739-1.571 | 0.698 | 1.068 | 0.718-1.588 | 0.746 |
| T3 (T3 ≥ 2.71) | 79/784 | 1.566 | 1.106-2.218 | 0.012 | 1.487 | 1.027-2.154 | 0.036 |
| Follow-up hsCRP |  |  |  |  |  |  |  |
| T1 (T1≤0.72) | 35/792 | 1 | Ref. |  | 1 | Ref. |  |
| T2 (0.73≤ T2≤1.54) | 32/792 | 0.906 | 0.561-1.463 | 0.685 | 0.913 | 0.557-1.499 | 0.719 |
| T3 (T3 ≥ 1.55) | 120/789 | 3.613 | 2.479-5.265 | <0.001 | 3.285 | 2.186-4.937 | <0.001 |
| Follow-up LDL-C |  |  |  |  |  |  |  |
| T1 (T1≤1.60) | 56/800 | 1 | Ref. |  | 1 | Ref. |  |
| T2 (1.60 < T2≤2.05) | 66/795 | 1.188 | 0.832-1.697 | 0.342 | 1.115 | 0.770-1.614 | 0.565 |
| T3 (T3 > 2.05) | 65/778 | 1.203 | 0.842-1.720 | 0.311 | 0.889 | 0.609-1.289 | 0.544 |

Major adverse cardiac and cerebrovascular events (MACCEs) included all-cause death, nonfatal MI, nonfatal stroke, rehospitalization for heart failure and any coronary artery revascularization.
The adjusted model included hypertension, diabetes mellitus, dyslipidemia, body mass index, previous PCI, CABG and MI, LVEF%, antiplatelet therapy, angiotensin blockade, diuretic and spironolactone use, multivessel disease, target vessel length, SYNTAX score and residual SYNTAX score. hsCRP, high-sensitive C reactive protein; LDL-C low-density lipoprotein cholesterol; HR, hazard ratio; CI confidence interval; PCI, percutaneous coronary intervention; CABG, coronary artery bypass grafting; MI, myocardial infarction; LVEF, left ventricular ejection fraction;

**Table 3.**
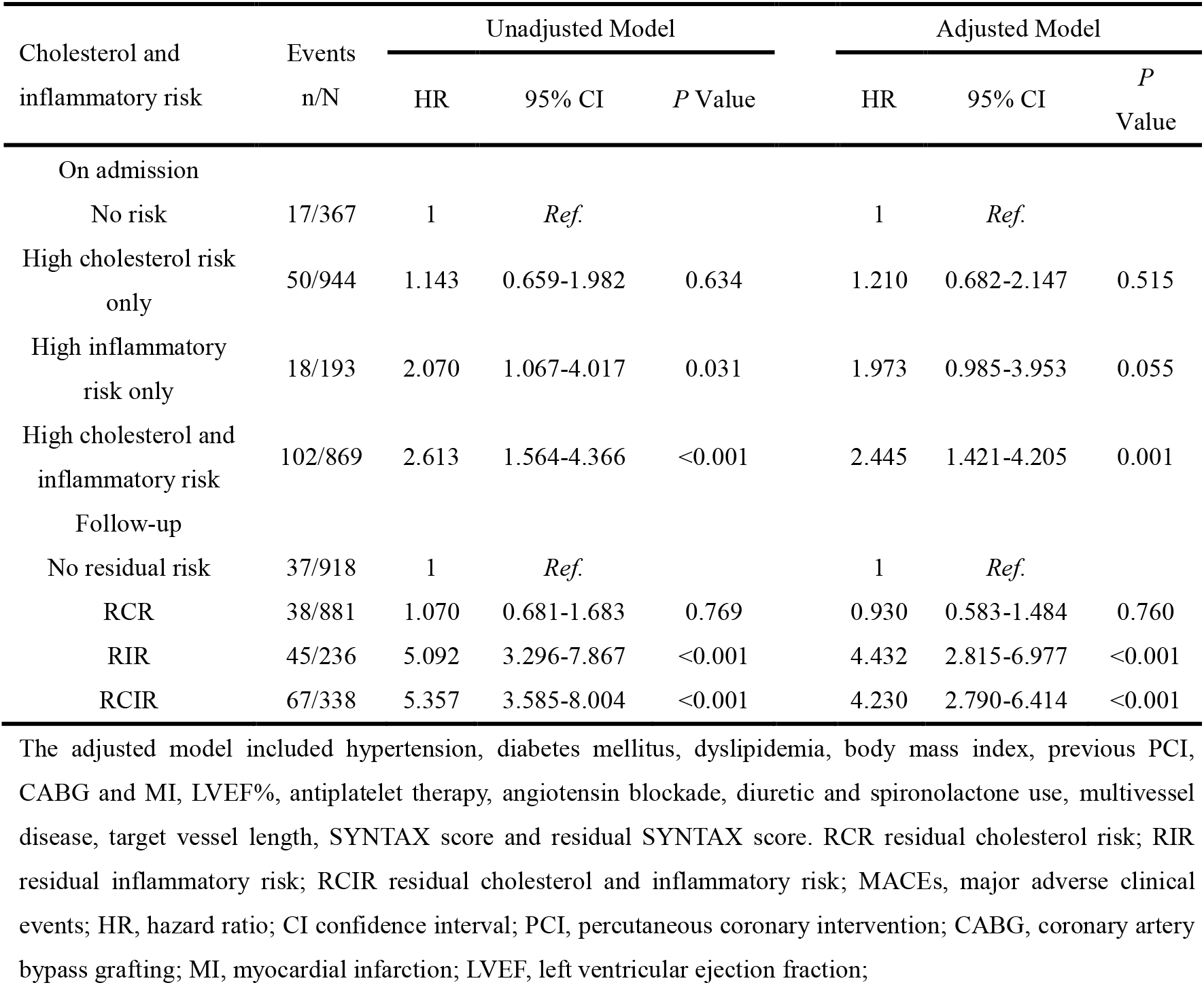
The predictive value of baseline and follow-up residual cholesterol and inflammatory risk for one-year MACCEs.

| Cholesterol and inflammatory risk | Events<br>n/N | Unadjusted Model |  |  | Adjusted Model |  |  |
| --- | --- | --- | --- | --- | --- | --- | --- |
|  |  | HR | 95% CI | *P* Value | HR | 95% CI | *P* Value |
| On admission |  |  |  |  |  |  |  |
| No risk | 17/367 | 1 | *Ref.* |  | 1 | *Ref.* |  |
| High cholesterol risk only | 50/944 | 1.143 | 0.659-1.982 | 0.634 | 1.210 | 0.682-2.147 | 0.515 |
| High inflammatory risk only | 18/193 | 2.070 | 1.067-4.017 | 0.031 | 1.973 | 0.985-3.953 | 0.055 |
| High cholesterol and inflammatory risk | 102/869 | 2.613 | 1.564-4.366 | <0.001 | 2.445 | 1.421-4.205 | 0.001 |
| Follow-up |  |  |  |  |  |  |  |
| No residual risk | 37/918 | 1 | *Ref.* |  | 1 | *Ref.* |  |
| RCR | 38/881 | 1.070 | 0.681-1.683 | 0.769 | 0.930 | 0.583-1.484 | 0.760 |
| RIR | 45/236 | 5.092 | 3.296-7.867 | <0.001 | 4.432 | 2.815-6.977 | <0.001 |
| RCIR | 67/338 | 5.357 | 3.585-8.004 | <0.001 | 4.230 | 2.790-6.414 | <0.001 |

The adjusted model included hypertension, diabetes mellitus, dyslipidemia, body mass index, previous PCI, CABG and MI, LVEF%, antiplatelet therapy, angiotensin blockade, diuretic and spironolactone use, multivessel disease, target vessel length, SYNTAX score and residual SYNTAX score. RCR residual cholesterol risk; RIR residual inflammatory risk; RCIR residual cholesterol and inflammatory risk; MACEs, major adverse clinical events; HR, hazard ratio; CI confidence interval; PCI, percutaneous coronary intervention; CABG, coronary artery bypass grafting; MI, myocardial infarction; LVEF, left ventricular ejection fraction;

## Discussion

The main findings of this study were as follows: 1) Almost half of PCI-treated patients still suffered from a high cholesterol risk burden despite contemporary lipid-lowering therapy. 2) On-admission and follow-up hs-CRP levels showed a consistent association with the risk of MACCEs, while the predictive value of LDL-C levels was attenuated during follow-up. 3) A combined effect of on-admission high cholesterol and inflammatory risk was found that could independently predict the prognosis of PCI-treated patients, while a single residual risk factor could not. 3) Inflammatory risk assessed by hs-CRP was a stronger predictor for the risk of MACCEs than cholesterol risk measured by LDL-C in PCI-treated patients after accepting contemporary statin therapy. 4) On-admission high inflammatory risk only could predict the risk of MACCEs independent of the cholesterol risk measured by LDL-C in patients with diabetes mellitus and accepting underpowered statin therapy. These data may have clinical implications for the following reasons: 1) Intensified lipid-lowering therapy after PCI treatment should still be recommended in current practice. 2) Anti-inflammatory therapy beyond traditional statin treatment should be considered after PCI treatment in patients with high inflammatory risk during follow-up. 3) It may be reasonable to initiate therapies targeting inflammation according to on-admission high inflammatory risk in diabetic patients accepting PCI treatment.

In the past decades, advancement in vascular biology has reshaped our understanding of atherosclerosis. It has shifted from the disease of lipid accumulation in arterial walls to the multifactorial and inflammatory-driven disease. In this novel perspective, inflammation and cholesterol both play similar roles in initiating and progressing atherosclerosis, through the dysregulation of endothelial dysfunction to promote LDL-C uptake, oxidation and accumulation to the erosion and rupture of the plaque, with the consequence of acute ischemic syndrome[20]. Statins remained the cornerstone therapy for the secondary prevention of patients with established cardiovascular diseases or multiple cardiovascular risk factors due to their pleiotropic effect in lowering cholesterol levels, stabilizing plaques, improving endothelial function and alleviating vascular inflammation[21]. Previous RCTs have substantiated the effectiveness of statin treatment in reducing recurrent cardiovascular events[2]. However, for advanced atherosclerotic cardiovascular disease patients, increased risks for recurrent events can still be foreseen during long-term follow-up despite an early coronary revascularization strategy and guideline-recommended medical therapy, an issue commonly described in clinical practice as the problem of ‘residual risk’[3, 22, 23]. Cholesterol undoubtedly represented a major residual risk factor and was defined as an unachieved LDL-C level goal despite current lipid-lowering therapy. While the precise goal for LDL-C remained in relentless debate, current clinical practice guidelines provide Class I recommendations for LDL-C targets of less than 1.8 mmol/L (70mg/dL) in most patients with atherosclerotic cardiovascular diseases[15]. The concept of ‘the lower the better’ for LDL-C levels has brought intensified lipid-lowering therapy into clinical practice. Aggressive lipid-lowering therapies such as intensified statin treatment or the use of ezetimibe and PCSK9 inhibitors on the basis of statins, have produced positive results and further reduced the risk of recurrent events by 2-15%[5–7]. In the current study, the enrolled participants were all PCI-treated patients, most of whom accepted moderate (74.3%) or more aggressive lipid-lowering treatment (20.4%) at discharge. However, only 25.7% of enrolled patients accepted GRST at discharge, and a high cholesterol risk burden (LDL-C≥1.8 mmol/L) could still be foreseen in almost half of enrolled patients (51.4%) after 1 to 3-month of follow-up, indicating the importance of intensified lipid-lowering therapies in current practice. Although the predictive value of high cholesterol risk measured by LDL-C for MACCEs was mediated by contemporary statin treatment in the current study, the beneficial effect of intensified lipid-lowering therapies could not be simply explained by the achieved LDL-C goal. A retrospective study was conducted in Asian patients who had either achieved LDL-C targets or received high-intensity statins with 1 year after PCI. The study revealed that compared with those who attained LDL-C goals without high-intensity statin treatment, those accepting high-intensity statins with or without LDL-C goal achievement were associated with a lower adjusted risk of major cardiovascular outcomes at 5 years, suggesting the importance of routine intensified lipid-lowering therapies after PCI treatment[24]. Further, a combined effect of on-admission cholesterol and inflammation risk for predicting the prognosis of PCI-treated patients was also found in the current study. The pleiotropic effects of intensified lipid-lowering therapies were also shown in the aspect of inhibiting inflammation. In the IMPROVE-IT trial, the rate of stabilized after acute coronary syndrome (ACS) patients with no residual cholesterol and inflammatory risk was significantly higher after statin adjunctive (simvastatin + ezetimibe) treatment than after simvastatin alone, and a better cardiovascular outcome was seen in those who achieved both hsCRP and LDL-C targets[25]. Considering the combined effect of cholesterol and inflammation on the prognosis of PCI-treated patients, intensified lipid-lowering therapy after PCI treatment remained necessary.

Another important determinant of residual risk in current practice is inflammatory risk as measured by hs-CRP. The concept of ‘dual targets of inflammatory and cholesterol risk’ has been confirmed in the IMPROVE-IT trial, with an improved clinical outcome in those patients with LDL-C<70 mg/dL and hs-CRP<2 mg/L during follow-up[25]. Post hoc analysis from large-scale clinical trials has substantiated that hs-CRP could still independently predict cardiovascular risk even in patients with extremely low LDL-C levels despite aggressive lipid-lowering therapies[22]. High inflammatory risk continued to persist after PCI treatment, ranging from 18.3% in East Asian populations to 38.0% in Western populations[9, 10]. Persistently high inflammatory risk was a reliable predictor for prognosis even in patients with baseline LDL-C<1.8 mmol/L, indicating that combination therapy with anti-inflammatory agents should be considered beyond lipid-lowering therapy for patients with high inflammatory risk[11]. In the current study, the rate of PCI-treated patients with persistent high inflammatory burden was 18.0%, which is consistent with previous findings in East Asian populations, showing that almost one-fifth of the PCI-treated populations were under persistent high inflammatory burden despite intensified statin therapy. Furthermore, the relative importance of inflammatory and cholesterol risk might have changed under the current lipid-lowering therapies. Recently, a collaborative analysis of three RCTs with 31245 established ASCVD patients, of which almost 50% were on high intensity statin treatment, unveiled significant insight. Among statin-treated individuals without high inflammatory risk, the unachieved goal of LDL-C did not seem to be associated with cardiovascular death or all-cause mortality. Instead, it was noted that hs-CRP, the biomarker for inflammation, emerged as a stronger predictor for the risk of future cardiovascular events and death compared to LDL-C among patients receiving contemporary statins[8]. Burgeoning evidence from large clinical trials of therapies targeting inflammation in atherosclerotic cardiovascular disease individuals is now emerging. It was not until 2017 with the publication of the CANTOS trial that proof of inflammation targets in atherosclerosis was provided. In the CANTOS trial, participants with a history of myocardial infarction and high inflammatory risk were randomly allocated to the treatment of canakinumab (an IL-1β inhibitor) or placebo group on the basis of standard medical therapy. Canakinumab lowered cardiovascular event rates by 15-17% compared with placebo without affecting LDL-C and apolipoprotein B levels, demonstrating that inhibition of the nucleotide-binding domain, leucine-risk-containing family, pyrin domain 3 (NLRP3) to interleukin-1 to interleukin-6 pathway was a crucial treatment target for atherosclerosis[26]. However, considering the cost effectiveness of canakinumab and the increased number of deaths related to infection, anti-inflammatory drugs with similar effects and no significant infectious side effects should be encouraged in current practice. Colchicine, a generic drug used to treat gout and Mediterranean fever, could suppress neutrophil deformability and extravasation and inhibits the intracellular transport of the caspase-recruitment domain, preventing localization of NLRP3 components and leading to reductions in interleukin-1β and interleukin-6. Reduction of the inflammatory burden with colchicine has emerged as a therapeutic option for secondary prevention in established coronary artery disease patients. In the COLCOT trial, patients after myocardial infarction were randomly assigned to treatment with colchicine 0.5 mg daily in comparison with placebo over a 2-year follow-up, with a 23% relative reduction in the primary endpoint[12]. Similar results were achieved in the LoDoCo2 trial, with a 31% risk reduction of the primary endpoint in chronic coronary disease patients[13]. In June 2023, the U.S. Food and Drug Administration approved the use of low-dose colchicine to reduce cardiovascular death, myocardial infarction, ischemic stroke or coronary revascularization in adult patients with established atherosclerotic disease or with multiple risk factors for cardiovascular diseases[27]. However, the widespread use of colchicine in coronary artery disease patients may cause concerns about the tendency toward a higher incidence of non-cardiovascular death and severe gastrointestinal adverse events among colchicine-treated patients[28]. Neutral result was also observed in a study concentrating on the effect of colchicine on PCI-related myocardial injury[29]. Hence, the administration of anti-inflammatory agents to a selected population in which a net clinical benefit could be achieved may be the most appropriate. In the current study, high inflammatory risk after accepting contemporary statin treatment was a major determinant of MACCEs independent of LDL-C level, indicating the necessity of initiating anti-inflammatory agents in that population. A noteworthy result found in this study was that on-admission high inflammatory risk only failed to independently predict the occurrence of MACCEs, making the timing of initiating anti-inflammatory therapy elusive. There may be several plausible explanations. First, the inflammation level can be dynamically changed over the early phase in unstable patients. A total of 68.3% of enrolled participants presented with ACS, and the inflammatory level could be stabilized after PCI treatment. In addition, high inflammatory risk on admission can also be alleviated by contemporary statin treatment at discharge. Hence, serial measurements of hs-CRP should be emphasized following PCI treatment to identify patients at high inflammatory risk. Recent guidelines about the use of colchicine among established cardiovascular disease patients recommend a daily use of 0.5 mg for patients with stable ischemic heart disease who, despite recommended therapy, have hs-CRP≥2 mg/L[27]. However, whether initiating anti-inflammatory agents during the early phase of acute ischemic heart disease according to high inflammatory risk still needs to be further explored. Furthermore, another important finding in the subgroup analysis was that on-admission inflammatory risk only could predict the risk of MACCEs independent of LDL-C in diabetic patients, suggesting the potential benefit of initiating therapies targeting inflammation earlier in diabetic patients undergoing PCI treatment. It is well known that diabetes mellitus is characterized by low-grade systemic inflammation. The underlying mechanism could be that hyperglycemia induce excessive reactive oxidative stress by the mitochondrial electron transport chain, thus accelerating the atherosclerotic process[30]. Hence, glucose-lowering therapies with anti-inflammatory effects could be the optimal strategy for diabetic patients with high inflammatory risk after PCI treatment. Recently, exploratory analysis has substantiated the effectiveness of semaglutide in reducing the inflammatory level ratio-to-baseline versus comparators in diabetes mellitus patients, potentially by a direct effect of semaglutide, independent of HbA1c or body weight reduction[31, 32]. The potential anti-inflammatory effect of semaglutide could be that semaglutide could effectively lower the level of CD163, a biomarker for macrophage activation, and reversal changes in leukocyte recruitment, rolling, and adhesion/extravasation related to the inflammatory response were also observed in animal models after semaglutide treatment[33]. Therefore, it may be reasonable to initiate glucose-lowering therapy such as semaglutide in diabetic patients with high inflammatory risk after PCI treatment to lower recurrent vascular events.

Several limitations should be acknowledged. First, this was a single-center observational retrospective cohort study in which many PCI-treated patients failed to complete a follow-up visit with measurements of hs-CRP and LDL-C, so potential selection bias invariably existed. Second, the enrolled population in this study underwent only one-year of follow-up with a relatively low MACCE rate, which may limit the statistical analysis and make it difficult to find a connection between on-admission and follow-up inflammatory and cholesterol risk with a single component of MACCEs. Finally, the statin dosage and duration information of the enrolled population before admission could not be obtained; hence, whether the change in statin intensity had an impact on the prognostic value of inflammatory and cholesterol risk still needs to be further explored.

## Conclusion

Anti-inflammatory agents should be considered in PCI-treated patients with high inflammatory risk after accepting contemporary statin therapy. Therapies targeting inflammation should be initiated early in diabetic patients with high inflammatory risk after PCI treatment.

## Data Availability

The datasets and materials mentioned above are available from the authors on reasonable requests

**Appendix**

## Acknowledgements

Not applicable

## Acronyms

PCI Percutaneous coronary intervention; CABG Coronary artery bypass grafting; HsCRP High-sensitive C-reactive protein; LDL-C Low-density lipoprotein cholesterol; HDL-C High-density lipoprotein cholesterol; MACCEs Major adverse cardiovascular and cerebrovascular events; GRST Guideline-recommended statin therapy; HR Hazard ratio; CI Confidence interval; RCR Residual cholesterol risk; RIR Residual inflammatory risk; RCIR Residual cholesterol and inflammatory risk; CAD Coronary artery disease; ACS Acute coronary syndrome; NSTEMI Non-ST segment elevated myocardial infarction; STEMI ST segment elevated myocardial infarction; SAP Stable angina pectoris; RCT Randomized controlled trial; PCSK9 Proprotein convertase subtilisin/kexin type 9; NLRP3 Nucleotide-binding domain, leucine-risk-containing family, pyrin domain 3; HbA1c Glycated hemoglobin; LVEF Left ventricular ejection fraction; SBP Systolic blood pressure; DBP Diastolic blood pressure; BMI Body mass index; IMPROVE-IT Improved Reduction of Outcomes: Vytorin Efficacy International Trial; FOURIER Further cardiovascular OUtcomes Research with PCSK9 Inhibition in subjects with Elevated Risk; SYNTAX SYNergy between PCI with TAXUS™ and Cardiac Surgery; CANTOS Canakinumab Anti-inflammatory Thrombosis Outcome Study; COLCOT Colchicine Cardiovascular Outcomes Trial; LoDoCo2 trial Low-dose colchicine2 trial;

## Availability of data and materials

The datasets and materials mentioned above are available from the authors on reasonable requests

## Ethics and consent to participate

This study has been approved by the ethics committee of Fuwai Hospital (No. 2021-1063). The informed consent from participants was waived by the Ethics Committee.

## Consent for publications

Not applicable

## Authors’ contributions

Conception and design of study: Ang Gao and Zifeng Qiu. Acquisition of data: Hong Qiu. Analysis and/or interpretation of data: Ang Gao and Zifeng Qiu. Drafting the manuscript: Ang Gao. Revising the manuscript critically for important intellectual content: Hong Qiu. Approval of the version of the manuscript to be published: Ang Gao, Zifeng Qiu, Yong Wang, Tingting Guo, Yanan Gao, Qianhong Lu, Zhiqiang Yang, Zhifan Li, Hong Qiu, Runlin Gao.

